# Large-scale psychometric assessment and validation of the Modified COVID-19 Yorkshire Rehabilitation Scale (C19-YRSm) patient-reported outcome measure for Long COVID or Post-COVID syndrome

**DOI:** 10.1101/2025.06.27.25330396

**Authors:** M Horton, AB Smith, S Halpin, R O’Connor, R Milne, D Winch, C Rayner, R Rocha Lawrence, DC Greenwood, ND Barkley, R Evans, J Kwon, H Dawes, C Wood, P Williams, H Master, M Mansoubi, G Mir, JH De Kock, J Mullard, M Ormerod, S Petrou, DB O’Connor, LOCOMOTION Consortium, M Sivan

**Affiliations:** Leeds Institute of Rheumatic and Musculoskeletal Medicine, University of Leeds, Leeds, UK; University of Southampton, Southampton, UK; Kent & Medway ICB Patient Reference Group, Kent, UK; University of Leeds LOCOMOTION Patient Advisory Group, UK; ELAROS 24/7 Ltd., UK; Leeds Institute for Data Analytics, University of Leeds, Leeds, UK; Northern Care Alliance, UK; Manchester Metropolitan University, Manchester, UK; Department of Respiratory Sciences, University of Leicester, UK; Nuffield Department of Primary Care Health Sciences, University of Oxford, Oxford; Medical School, NIHR Exeter Biomedical Research Centre, Faculty of Health and Life Sciences, University of Exeter, Exeter, UK; Birmingham Community Healthcare NHS Foundation Trust, UK; Hertfordshire Community NHS Trust, UK; School of Medicine, University of Leeds, UK; NHS Highland COVID Recovery Service, North West University, Faculty of Health Sciences, UK; Faculty of Medical Sciences, Newcastle University, UK; Long Covid Support (UK Registered Charity: 1198938); School of Psychology, University of Leeds, UK; COVID Rehabilitation Service, Leeds Community Healthcare NHS Trust, Leeds, UK; National Demonstration Centre of Rehabilitation Medicine, Leeds Teaching Hospitals NHS Trust, Leeds, UK

**Keywords:** Post-COVID Syndrome, Long COVID, C19-YRSm, patient-reported outcome

## Abstract

**Background:** The C19-YRS was the first condition-specific scale for Long COVID/Post-COVID syndrome. Although the original C19-YRS evolved to the modified version (C19-YRSm) based on psychometric evidence, clinical content relevance and feedback from patients and healthcare professionals, it has not been validated through Rasch analysis.

**Objectives:** To psychometrically assess and validate the C19-YRSm using newly collected data from a large-scale, multi-centre study (LOCOMOTION).

**Methods:** 1278 patients (67% Female; mean age = 48.6, SD 12.7) digitally completed the C19-YRSm. The psychometric properties of the C19-YRSm Symptom Severity (SS) and Functional Disability (FD) subscales were assessed using a Rasch Measurement Theory framework, assessing for individual item model fit, targeting, internal consistency reliability, unidimensionality, local dependency (LD), response category functioning and differential item functioning (DIF) by age group, sex and ethnicity.

**Results:** Rasch analysis revealed robust psychometric properties of both subscales, with each demonstrating unidimensionality, appropriate response category structuring, no floor or ceiling effects, and minimal LD and DIF. Both subscales also displayed good targeting and reliability (SS: Person Separation Index (PSI)=0.81, Cronbach’s alpha=0.82; FD: PSI=0.76, Cronbach’s alpha=0.81).

**Conclusion:** Although some minor anomalies are apparent, the modifications to the original C19-YRS have strengthened its measurement characteristics, and its clinical and conceptual relevance.

## Introduction

Long Covid (LC) or Post COVID-19 Syndrome (PCS) is a multisystem clinical syndrome where symptoms persist for more than three months after acute infection with SARS-CoV-2. ^1^ The prevalence of the condition is estimated to be approximately 1.8 million cases in the UK alone^2^ and 200 million individuals worldwide. ^3^

Common symptoms include fatigue, shortness of breath, cognitive impairment, muscle and joint pain, chest pain, palpitations, persistent loss of smell and taste, gastro-intestinal upset, and headache. ^4^ Symptoms may fluctuate^5^ with approximately 20% of those suffering with LC describing their symptoms as severe. ^2^ For some individuals, symptoms may persist for more than 4 years (persistent LC) following the initial COVID-19 infection. ^6^ This protracted course of LC leads to a significant compromise on individuals’ ability to work and conduct day-to-day tasks and can result in severely reduced health-related quality of life. ^7-9^

Measuring symptom burden, functional disability (or ability) and health-related quality of life (HRQoL) through patient-reported outcome measures (PROMs) is therefore crucial to understand the impacts on health, condition trajectories and the cost-effectiveness of interventions.

The COVID-19 Yorkshire Rehabilitation Scale (C19-YRS) is an LC condition-specific PROM. The instrument has been widely employed in primary care and community settings, ^10-12^ rehabilitation interventions, ^13^ and post-COVID epidemiological studies. ^14,15^ Following an initial psychometric analysis of the C19-YRS, ^16,17^ psychometric and clinical evidence, and feedback from patients and healthcare professionals was subsequently integrated, culminating in a modified version of the instrument (C19-YRSm). ^18^ The C19-YRSm has since undergone further classical psychometric validation and has been shown, for instance, to have good internal reliability and convergent validity in a Croatian patient population. ^19^ More recent validation has demonstrated the C19-YRSm to have good psychometric properties, ^20^ in terms of internal consistency and test-retest reliability, as well as discriminant and convergent validity. Factor analysis supported the instrument’s factor structure. Furthermore, an exploratory minimal important difference (MID) and minimal clinically important difference (MCID) were determined for the subscale scores. ^20^

However, the psychometric properties of the modified C19-YRSm have not yet been assessed using Rasch measurement theory (RMT). In contrast to classical psychometrics, which are focused on the test-level, i.e., the instrument as a whole, RMT allows for item-level analysis. This enables the identification of individual PROM items that may potentially require modification (or removal) in order to improve the measurement properties of a PROM. The aim of this study was therefore to use Rasch measurement methodology to psychometrically assess and further validate the C19-YRSm using data collected from a large-scale, multi-centre study (LOCOMOTION). ^21^

## Materials and Methods

### C19-YRSm

The C19-YRSm consists of four separate subscales: Symptom Severity (SS), Functional Disability (FD), Other Symptoms (OS), and Overall Health (OH). The OH subscale is a single item, scored on a 0-10 numeric rating subscale, with a score of 0 representing “worst health” and 10 being “best health”. Given the OH is a single-item subscale it cannot be analysed using the Rasch model. The OS subscale consists of a checklist of 25 additional symptoms, where respondents select the symptoms that they have experienced over the last 7 days based on yes/no options. The analysis of the OS subscale is not presented here.

For the remaining subscales, both the SS (26 items condensed to 10 core items, see below) and FD (5 items) are summed individually to form total scores for each subscale. All items on the SS and FD subscales are scored on a 4-point subscale (0=No problem; 1=Mild problem; 2=Moderate problem; 3=Severe problem) where a higher score represents a higher severity of the problem, i.e. worse symptoms or worse functional disability.

Some of the SS subscale core items are grouped within subsets. For these items, the maximum value observed within the subset is selected as the representative value. This scoring step is taken due to the inherent (local) dependency between the items within a subset, as observed during the modification of the original C19-YRS. ^18^ Taking the maximum score from within a set avoids local dependency, whilst maintaining the clinical utility of the individual component items. The sections concerning breathlessness, cough/throat sensitivity, smell/taste, pain/discomfort, cognition, palpitation/dizziness, and anxiety/mood each have a maximum score that is taken from across multiple items in the section.

### Data Collection

Data collection was carried out as part of the LOCOMOTION study^21^ (NIHR Ref: COV-LT2-0016), with the C19-YRSm data collected routinely within 10 participating Long Covid services across the UK between December 2021 and October 2023. Consent and clinical data were collected using the ELAROS digital patient-reported outcome measures platform. ^22^. Ethics approval for the LOCOMOTION study was obtained from the Bradford and Leeds Research Ethics Committee on behalf of Health Research Authority and Health and Care Research Wales (reference: 21/YH/0276).

### Rasch analysis

Rasch analysis of the data^23^ was completed with RUMM2030 software, ^24^ and carried out separately for the SS subscale (10 items) and the FD subscale (5 items). Key criteria for the RMT are 1) Unidimensionality - whether the items represent a single factor, 2) Item fit - whether the items fit the Rasch model, 3) Local Dependence - the absence of any further association between items beyond that explained by the underlying trait, 4) Response category functioning (or threshold disordering) - requiring the latent trait to increase monotonically across item response categories and 5) Item invariance (or absence of item bias or differential item functioning (DIF)) - requiring item properties to be invariant to subgroup characteristics (e.g., gender, ethnicity) where latent trait levels are equivalent.

1. Unidimensionality was evaluated by a series of t-tests, ^25^ with multidimensionality indicated if independent subsets of items delivered significantly different person estimates, and the lower bound 95% CI percentage of significantly different t-tests was > 5%.
2. The Rasch analytic process included several standard tests of fit, covering both the overall subscale and item-level fit. All items were assessed individually for fit to the Rasch model (Partial Credit Model) ^26^ within the subscale item set to assess whether each item contributes to the underlying construct. Item misfit was indicated where the Bonferroni-adjusted chi-squared p-value was statistically significant for an item and the standardised (z-score) fit-residuals fell outside ±2.5. ^27,28^
3. Tests of local dependency (LD) were carried out to determine whether any items in the subscale were more closely related than is explained by the underlying construct; LD was indicated using a residual correlation (Q3 value) criterion cut point of 0.2 above average residual correlation. ^29^
4. Response category functioning was assessed to determine whether the response structure of the items was operating in the intended manner. A functional 0-3 response category structure for each item would be indicated by sequential response thresholds (the crossover points between subsequent response categories) on the underlying (logit) subscale. ^30^
5. Item bias was assessed through uniform and non-uniform differential item functioning (DIF) testing by sex (male/female), age group (16-49, 50plus) and ethnicity, with significant DIF indicated at a Bonferroni-adjusted ANOVA p-value.

Furthermore, reliability indices were taken as the person separation index (PSI), and the Cronbach’s alpha values, and the scale-sample targeting was assessed graphically through the relative distribution of item and person locations, along with the calculation of floor and ceiling effects. When the Rasch model assumptions are satisfied, the sufficiency of the raw score allows for the transformation into a linear, interval-level transformation. ^28^

### Cross-validation

In order to assess the replication of results across independent samples, the complete sample was randomly split into three equally-sized subsamples which were examined separately. This strengthens the analysis through replication, whilst avoiding the over-powering of RUMM fit statistics and misinterpretation that can occur with sample sizes >500. ^31^ The subsamples were used to assess Rasch-based individual item fit and DIF. However, the complete sample was utilised to assess response category functioning, targeting, local dependency and the reliability indices, as these tests operate better with the precision afforded by larger sample sizes. To allow for brevity of reporting, only the results of the first subsample are presented within the manuscript.

### Working group/Patient Advisory Group

All empirical results were reported back to a working group made up of clinicians, patients, and social scientists with expertise in PROMs and psychometrics, and additionally to the wider LOCOMOTION team, for sense-checking from both the patient and clinical context. Results were also reported back to a patient advisory group (PAG), and any potential further modifications were discussed within the working group and the PAG, with an emphasis on the practical implications of any potential change in the instrument.

## Results

### Sample

Data from 1278 patients were included in the study. The mean age was 48.6 (standard deviation, SD: 12.7) years, predominantly female (67%) and White (79%). The key demographic characteristics of the sample are presented in Table 1. The data for the presented cross-validation subsample were from 423 patients.

**Table 1.**
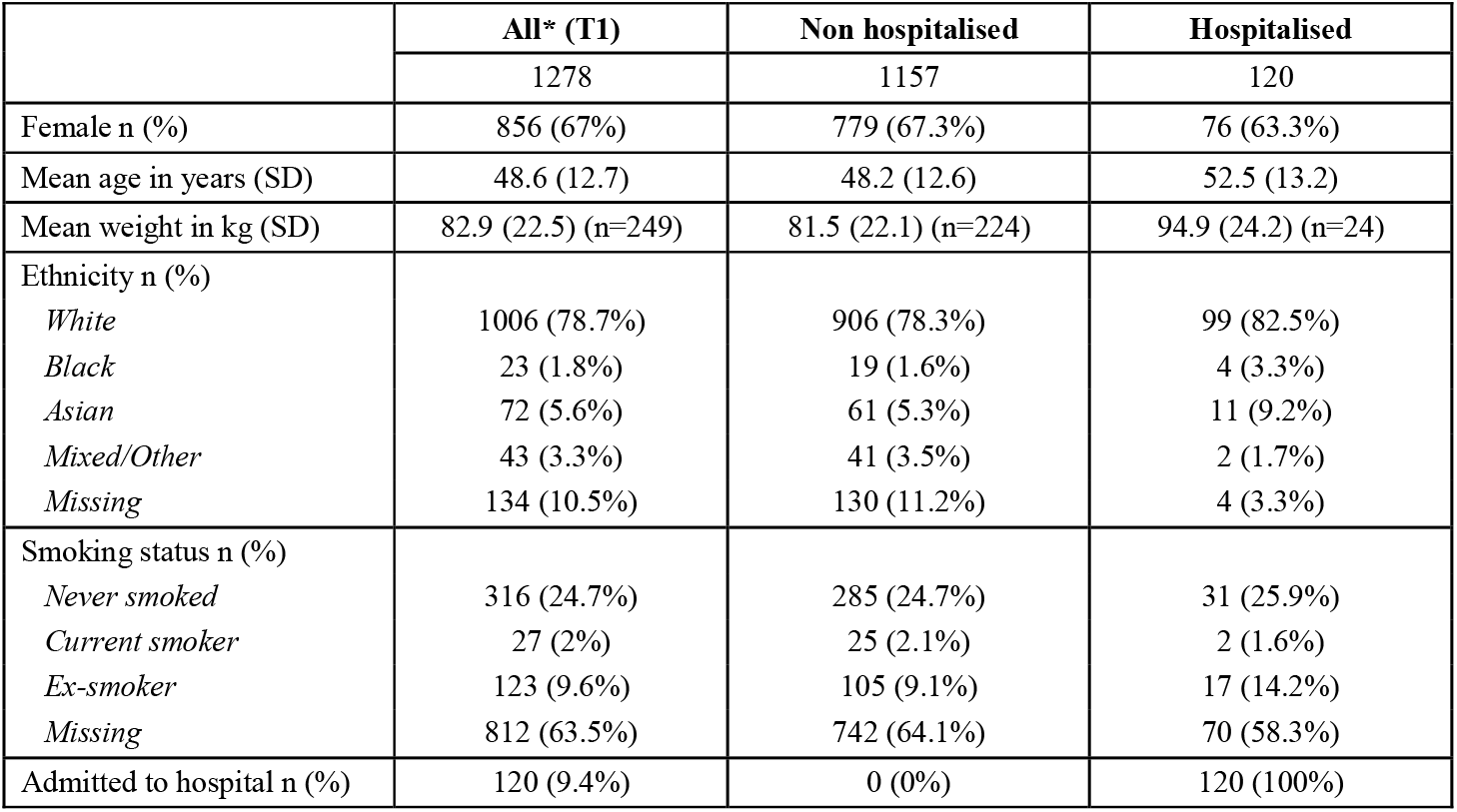
Demographic characteristics of sample.

### Rasch Analysis

#### Symptom Severity Subscale: Unidimensionality, reliability, floor/ceiling and targeting

A summary of the psychometric properties of both subscales are presented in Table 2, including results for the full sample and for the cross-validation subsample. The results indicated that the SS subscale was unidimensional, although the percentage of significant t-tests fell just outside the lower bound 95% CI, 5% criterion (5.1%) on the full sample. A good level of internal-consistency reliability (0.82) was displayed, with no floor or ceiling effect (see Table 2, and Figure 1).

**Table 2.** Overview of psychometric properties of the Symptom Severity (SS) and Functional Disability (FD) subscales.

|  | Symptom Severity (SS) |  | Functional Disability (FD) |  |  |
| --- | --- | --- | --- | --- | --- |
|  | Full Sample | Sample A | Full Sample | Sample A | Target Values |
| total n - (extremes) = valid n | **1268 - (11) = 1257** | **423 - (3) = 420** | **1268 - (63) = 1205** | **423 - (17) = 406** |  |
| Number of items | **10** | **10** | **5** | **5** |  |
| Overall Scale Fit (Chi-square p) | **p<0.001** | **p<0.001** | **p<0.001** | **p=0.026** | p>0.01 |
| Individual Item Fit Residuals | 6/10 out of range | 2/10 out of range | 2/5 out of range | 1/5 out of range | within +/-2.5 |
| Individual Item Fit (Chi-square p) | 7/10 out of range | 4/10 out of range | 3/5 out of range | All in range | p>0.05 (Bonferroni adj) |
| Person Fit Residuals | 1.4% outside range | 1.2% outside range | 1.3% outside range | 1.2% outside range | within +/-3.0 |
| Unidimensionality | 6.28% (5.1%) | 6.43% (4.3%) | 2.66% | 2.46% | % of significant t-tests <5% |
| Local Dependency | 2/45 pairwise significant | 2/45 pairwise significant | 1/10 pairwise significant | 1/10 pairwise significant | <0.2 above average Q3 |
| Targeting | good alignment | good alignment | good alignment | good alignment | aligned |
| % sample at floor (min score) | 0.0% | 0.0% | 3.0% | 3.3% |  |
| % sample at ceiling (max score) | 0.9% | 0.7% | 2.0% | 0.7% |  |
| % sample at floor and ceiling (combined) | 0.9% | 0.7% | 5.0% | 4.0% | <15% |
| Person Separation Index (PSI) | 0.81 | 0.82 | 0.76 | 0.77 | >0.85 |
| Cronbach's Alpha | 0.82 | 0.84 | 0.81 | 0.81 | >0.85 |
| Response category threshold ordering | 4/10 disordered | 2/10 disordered | All ordered | 3/5 disordered | All ordered |
| DIF-by-Sex | No DIF | No DIF | 1/5 display DIF (nu) | 1/5 display DIF (nu) | p>0.05 (Bonferroni adj) |
| DIF-by-Age Group | 3/10 display DIF | No DIF | 3/5 display DIF | No DIF | p>0.05 (Bonferroni adj) |
| DIF-by-Ethnicity | No DIF | No DIF | No DIF | No DIF | p>0.05 (Bonferroni adj) |

**Figure 1.**
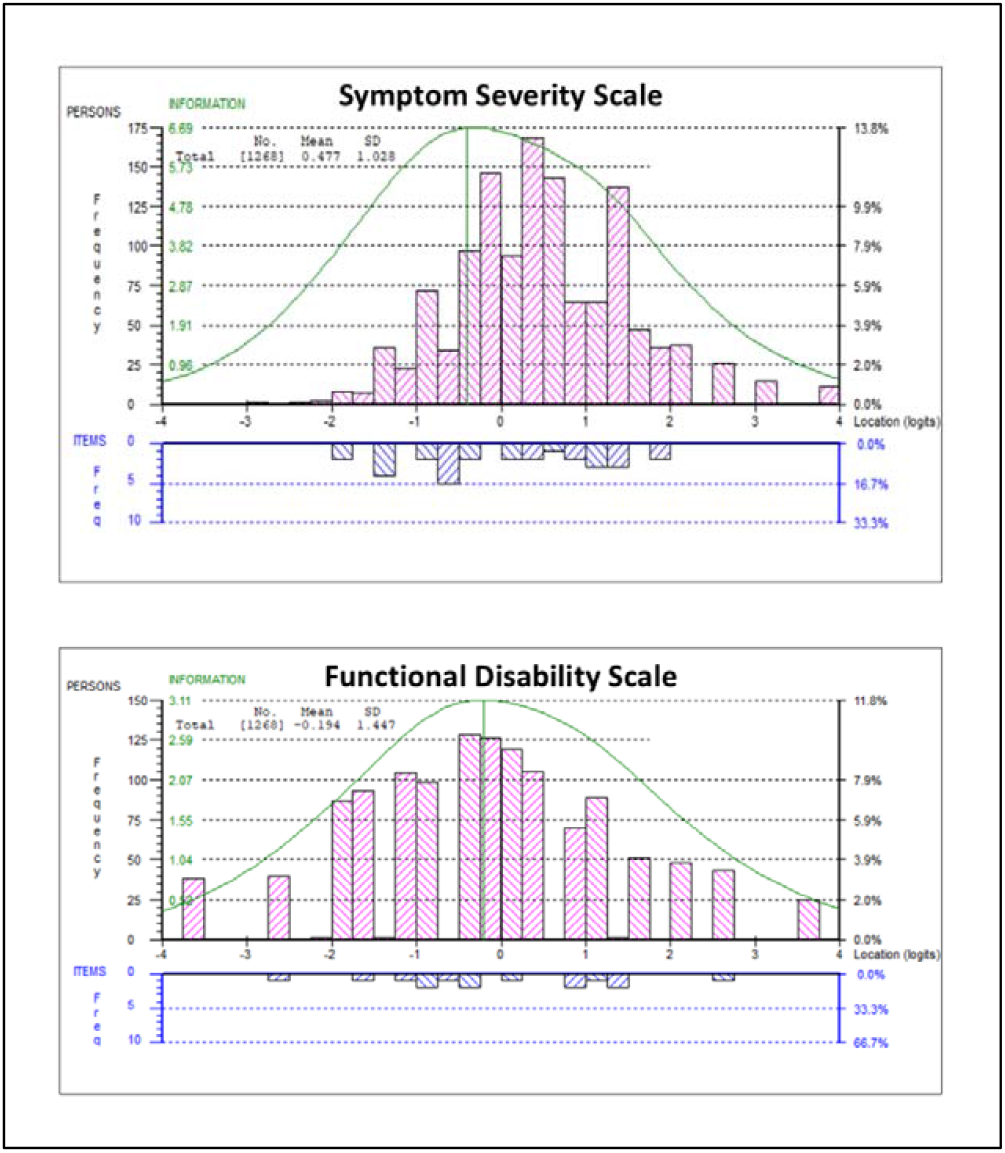
The relative scale-to-sample targeting of the Symptom Severity (SS) and Functional Disability (FD) subscales. The (logit) location distribution of the sample is plotted above the x-axis, and the (logit) location distribution of the scale items is plotted below the x-axis. Here, we can see a slight right-skew for the SS scale, and good targeting for the FD scale.

#### Symptom Severity Subscale: Local Dependency

The average residual correlation was -0.10 and therefore the criterion value to indicate LD was taken as 0.10 (−0.10 + 0.2). Two local dependencies out of the 45 (4%) pairs were identified (Table 3). The largest LD was observed between the “Fatigue” and “Post-exertional malaise” items (0.17); the other between the “Breathlessness” and “Cough” (0.13). Both of these dependencies appear to be conceptually logical, suggesting that these findings are real, and not just a chance finding.

**Table 3.**
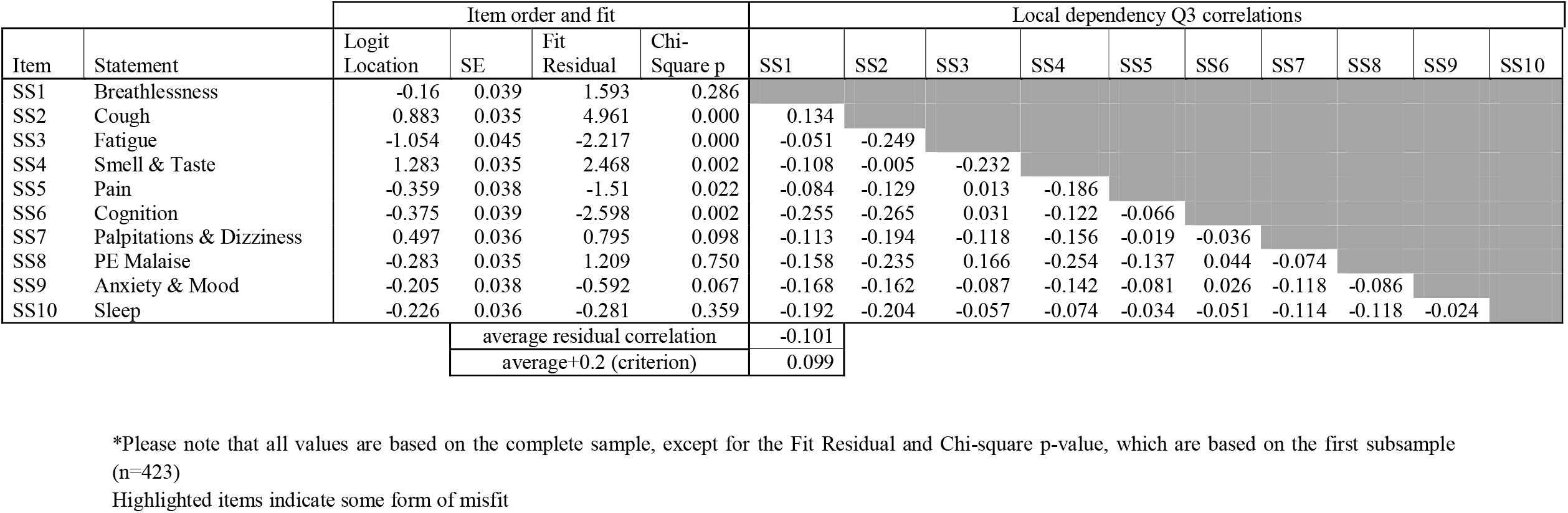
Individual item fit and local dependency of the Symptom Severity (SS) subscale.

#### Symptom Severity Subscale: Individual Item Fit

Most items were within the acceptable fit ranges, although misfit was identified for some items (Table 3). The “Cough” item (covering “cough” and “throat sensitivity”) displayed the largest chi-square and fit residual misfit anomalies, with the high positive fit residual value indicating an underdiscrimination. The “Smell/Taste” and “Cognition” items also demonstrated some misfit although this was borderline in both instances, and inconsistent among the different subsamples.

#### Symptom Severity Subscale: Category Response structure

The modified 4-response category format mostly displayed an ordered, functional response structure across all items, evidencing a marked improvement from the response functioning of the original C19- YRS. However, the ‘Smell/Taste’ item was consistently disordered among the full sample and all subsamples, with a non-borderline response structure suggesting that a binary response format may be more appropriate. Additionally, the full sample displayed three further items as borderline disordered (‘Fatigue’, ‘PEM’ and ‘Sleep’), with these same items either ordered or borderline disordered among the three smaller subsamples, suggesting that this is not problematic.

#### Symptom Severity Subscale: Item location ordering (easiest/most difficult items to affirm)

The item location ordering can be observed in Table 3. The “Fatigue” item marked the lowest location on the subscale, meaning that fatigue is observed as the most frequently problematic issue on the SS subscale, i.e., most easily endorsed item by people with LC. Conversely, the “Smell/Taste” item had the highest location on the subscale, meaning that smell/taste is observed as the least frequently problematic issue on the SS subscale.

#### Symptom Severity Subscale: Differential Item Functioning

No items displayed any consistent significant DIF by sex, age group or ethnicity grouping. However, the limited sample for minority ethnic groups was insufficient to make the finding on ethnicity reliable.

#### Symptom Severity Subscale: Post hoc analysis addressing the issues found

In order to determine whether amendments could be made to address and thereby potentially resolve the individual item issues that had been identified, the first subsample (N=423) was taken as an experimental dataset and a number of analysis iterations were run.

The first analysis focused on retaining all items in the SS subscale and involved rescoring the “Smell/Taste” and “Cough” items into a binary response format. This resolved the associated response structure and item fit issues. Furthermore, the “Breathlessness” and “Cough” items were subtested, i.e., added together into a single item, rather than contributing as two separate items, to account for local dependency. This resolved the dependency issues, although the subtested item displayed some misfit (Fit Residual: 3.8).

The second analysis focused on model fit and involved removing “Cough” whilst retaining the rescored (binary) “Smell/Taste” item. This resolved all fit and dependency issues, although the PEM item continued to display a borderline disordered response structure.

Finally, in order to examine the impact of these amendments on person estimates (“scores”), the person estimates from the complete subscale (full sample) were correlated against the full-sample person estimates from both analyses. ^32^ This indicated that there was strong (Spearman’s) correlation between the complete subscale person estimates and both post-hoc analyses estimates (0.99 and 0.98, respectively), indicating that the subscale amendments have very little effect on person ordering.

Taken together, this suggests that, despite the issues identified, it is perhaps optimal to retain the complete SS subscale in its original format, in order to retain maximum information and allow for continuity of data collection and comparison. Given that Rasch model assumptions have been satisfied, the transformation of the raw ordinal scale scores into interval-level equivalent scores is appropriate, and these transformed scores are available in Table 5. Please note that this transformation is only valid for complete data, where all items have been included in the total score.

#### Functional Disability subscale: Unidimensionality, reliability, floor/ceiling and targeting

A summary of the psychometric properties for the FD subscale is presented in Table 2. The FD subscale was unidimensional (only 2.6% of unidimensionality t-tests were statistically significant), displayed a good level of internal-consistency reliability (0.82), and had good subscale-sample targeting with no floor or ceiling effect (see Table 2 and Figure 1).

#### Functional Disability subscale: Local Dependency

The average correlation was -0.23 and therefore the criterion value to indicate LD was taken as -0.03 (−0.23 + 0.2). One local dependency (Table 4) was observed between the “Walking or moving around” and “Personal care” items. Again, there appears to be a conceptual connection between these two items, suggesting that this is a real dependency rather than a chance finding.

**Table 4.** Individual item fit and local dependency of the Functional Disability (FD) subscale.

| Item | Statement | Item order and fit |  |  |  | Local dependency Q3 correlations |  |  |  |  |
| --- | --- | --- | --- | --- | --- | --- | --- | --- | --- | --- |
|  |  | Logit Location | SE | Fit Residual | Chi-Square p | FD1 | FD2 | FD3 | FD4 | FD5 |
| FD1 | Communication | 0.035 | 0.039 | 3.54 | 0.010 |  |  |  |  |  |
| FD2 | Walking | -0.045 | 0.039 | 0.207 | 0.572 | -0.408 |  |  |  |  |
| FD3 | Personal Care | 1.459 | 0.044 | -0.672 | 0.599 | -0.384 | 0.017 |  |  |  |
| FD4 | Daily Living | -1.366 | 0.043 | -2.416 | 0.162 | -0.357 | -0.134 | -0.112 |  |  |
| FD5 | Social | -0.083 | 0.038 | -0.786 | 0.451 | -0.133 | -0.394 | -0.263 | -0.112 |  |
| average residual correlation |  |  |  |  |  | -0.228 |  |  |  |  |
| average+0.2 (criterion) |  |  |  |  |  | -0.028 |  |  |  |  |

\*Please note that all values are based on the complete sample, except for the Fit Residual and Chi-square p-value, which are based on the first subsample (n=423)
Highlighted items indicate some form of misfit

**Table 5.** The raw-score to interval-level equivalent transformed score for the Symptom Severity (SS) and Functional Disability (FD) subscales. *Please note that this conversion is only valid in the case of complete data

| Raw score | Interval transformations |  |
| --- | --- | --- |
|  | SS (0-30) | FD (0-15) |
| 0 | 0.00 | 0.00 |
| 1 | 2.80 | 1.84 |
| 2 | 4.74 | 3.19 |
| 3 | 6.07 | 4.17 |
| 4 | 7.13 | 4.97 |
| 5 | 8.03 | 5.67 |
| 6 | 8.81 | 6.32 |
| 7 | 9.53 | 6.96 |
| 8 | 10.19 | 7.61 |
| 9 | 10.82 | 8.28 |
| 10 | 11.41 | 8.98 |
| 11 | 11.99 | 9.72 |
| 12 | 12.57 | 10.55 |
| 13 | 13.14 | 11.56 |
| 14 | 13.71 | 12.99 |
| 15 | 14.29 | 15.00 |
| 16 | 14.87 |  |
| 17 | 15.47 |  |
| 18 | 16.09 |  |
| 19 | 16.72 |  |
| 20 | 17.36 |  |
| 21 | 18.02 |  |
| 22 | 18.70 |  |
| 23 | 19.40 |  |
| 24 | 20.16 |  |
| 25 | 20.98 |  |
| 26 | 21.92 |  |
| 27 | 23.06 |  |
| 28 | 24.55 |  |
| 29 | 26.75 |  |
| 30 | 30.00 |  |

#### Functional Disability subscale: Individual Item Fit

The “Communication” item consistently displayed a fit residual misfit anomaly with the high positive fit residual value indicating an underdiscrimination (Table 4). No other items indicated any evidence of misfit.

#### Functional Disability subscale: Category Response structure

The response categories displayed an ordered, functional response structure across all items. None of the items were disordered.

#### Functional Disability subscale: Item location ordering (easiest/most difficult items to affirm)

The “Other activities of daily living” item marked the lowest location on the subscale, i.e., the most frequently problematic (or easily endorsed) issue on the FD subscale. Conversely, the “Personal care” item had the highest location on the subscale, representing the least frequently problematic issue on the FD subscale. The item location ordering can be observed in Table 4.

#### Functional Disability subscale: Differential Item Functioning

There was no significant DIF by sex, age group or ethnicity grouping indicated for any of the items across any of the samples. As highlighted earlier, however, the ethnic minority sample was insufficiently powered to make the finding on ethnicity reliable. DIF by age group was observed for the “Walking or moving around” item, this was, however, consistent with expectation, as the older group are more likely to report issues with walking.

#### Functional Disability Subscale: Post hoc analysis addressing the issues found

Although there are relatively few issues found in the FD subscale, the first subsample (N=423) was again taken as an experimental dataset and amendments were made to address the issues that had been identified. This involved sub-testing the “Walking or moving around” and “Personal care” items in order to resolve the dependency issues, although this resulted in a further borderline dependency between the “Other activities of daily living” and “Social role” in the full sample. However, no further sub-testing was carried out, due to the borderline nature of the apparent dependency.

In order to examine the impact of this amendment on person estimates, the person estimates from the complete scale (full sample) were correlated against the full-sample person estimates from the resolved analysis. This indicated a very strong (Spearman’s) correlation of 0.999, indicating that the subscale amendment had very little effect on the ordering of persons. Again, this suggests that the parsimonious retention of the complete FD subscale in its original format would retain maximum information and allow for continuity of data collection and comparison.

Again, given that Rasch model assumptions have been satisfied, the transformation of the raw ordinal scale scores into interval-level equivalent scores is appropriate, and these transformed scores are available in Table 5 (for complete data).

### Working group/PAG

A benefit of the C19-YRSm is that it is a relatively short measure with a simple response structure. The feedback from the PAG suggested that it was not burdensome for patients to complete, and that the simplified response format was more appropriate than the previous 11-point numeric rating scale on the original C19-YRS. The PAG suggested that the C19-YRSm was comprehensible, easy to understand, and that the range of symptoms covered across the SS and OS scales (not presented here) was broadly comprehensive, whilst remaining manageable.

## Discussion

The aim of the study was to further validate the C19-YRSm following the initial development^17^ and subsequent psychometric analysis. ^18^ In line with the classical psychometric analysis, ^20^ the results demonstrated evidence of a two-factor structure, comprising unidimensional Symptom Severity and Functional Disability subscales, with both displaying good internal reliability.

Within the subscale analysis, the few item anomalies observed concerned the “Cough” and “Smell/Taste” items, both within the Symptom Severity subscale. It is uncertain as to why these items were not consistent with the other subscale items, but there are some potential explanations. For instance, these items also marked the ‘most difficult’ end of the subscale, meaning that they were generally reported to be problematic less frequently than the other items. This positioning means that there is less certainty in respect of the item fit characteristics of the items (given that less sample information is available). Furthermore, it also means that these items are an important demarcation of the upper measurement range of the subscale. The post-hoc removal and amendment of these items had very little effect on the ordering of the person estimates that were generated, therefore the added clinical and measurement information provided by the retention of these items would seem to outweigh any potential improvement in subscale fit. ^33^

Although the cough, throat sensitivity and anosmia items were recognised by the Working Group as common and important symptoms of Covid, the analysis results indicate that their contribution to the impact of Long Covid on a person’s daily life is less clear. It is also possible that a binary (no problem/problem) response format may be more appropriate for these items than the 4-point response structure, especially for the “Smell/Taste” item. Feedback from the Working Group suggested that a binary response would align with the manifestation of these symptoms that present in LC/PCS clinics, especially for the “Smell/Taste” item.

The input from the PAG also suggested that the C19-YRSm was not burdensome for patients to complete, and that the simplified response format was more appropriate than the previous 11-point numeric rating subscale. The benefit of the C19-YRSm is that it is a relatively short measure with a simple response structure. If outcome measures are to be repeatedly used in clinical or research settings, they should be relevant, clinically useful, and non-burdensome to patients. Long PROMs may lead to questionnaire response burden, which is recognised as a threat to subscale completion and adherence in trials. ^34^

This study is the first to the authors knowledge to investigate item bias or DIF for the C19-YRSm and demonstrated little or no DIF across age groups and gender, thus reinforcing the instrument usability across a wide population of people living with LC. However, one potential limitation to be noted here is that minority ethnic groups were considerably underrepresented within our patient sample. Although there was no evident item bias, there is nevertheless a need for further research that is sufficiently powered to explore any ethnic variation in LC symptoms.

## Conclusion

The modified C19-YRS has been demonstrated to have significant advantages over the original C19- YRS. The content coverage is much improved, including a number of common symptoms that were not included in the original version. This study and preceding validation studies have shown the Symptom Severity and Functional Disability subscales to be far more robust than the original C19- YRS, thereby strengthening the measurement characteristics of the C19-YRSm, along with enhancing the clinical and conceptual relevance for use in both clinical and community settings. For research purposes, an interval-level score transformation is available, allowing for the calculation of parametric statistics on C19-YRSm scores.

## Appendix

The C19-YRSm is available via the University of Leeds licensing platform, here: https://licensing.leeds.ac.uk/product/c19-yrs-covid-19-yorkshire-rehabilitation-subscale

## Declarations

### Ethics approval and consent to participate

Ethics approval for the LOCOMOTION study was obtained from the Bradford and Leeds Research Ethics Committee on behalf of Health Research Authority and Health and Care Research Wales (reference: 21/YH/0276; Trial registration number NCT05057260, ISRCTN15022307).

### Data availability statement

The datasets used and analysed during the current study are available from the corresponding author on reasonable request.

### Author contribution statement

MH, SP, RJOC, HD, RM, SH, GM, CR, DOC, DCG, NDB, MS were responsible for the funding acquisition. MH was additionally responsible for the data analysis and methodology; DCG and ABS additionally contributed to the methodology. All authors contributed to the drafting, review and editing of the manuscript.

## Acknowledgements

**-**

